# A Cross-Sectional Survey of Knowledge, Attitudes and Practices (KAP) towards COVID-19 in working professionals of Islamabad during lockdown

**DOI:** 10.1101/2024.11.24.24317870

**Authors:** Nadia Nisar, Murtaza Haider Haider, Aashifa Yaqoob, Inayah Safdar, Nazish Badar

## Abstract

**Background:** The first confirmed case of COVID-19 in Pakistan was reported on 26 February 2020. Along with several other regions, Islamabad was affected by COVID-19 epidemic. Smart lockdown was declared on 23 March 2020 in Islamabad. The purpose of the survey was to study the status of Knowledge, Attitude and Practice regarding COVID-19 among the government employees, residing in Islamabad city, during COVID-19 outbreak.

**Methods:** A week during smart lockdown, a cross sectional survey was conducted in Islamabad city between 5th to 19^th^ May 2020. Sociodemographic of participants were compared with one way, t-test, or Chi-square test. Data analysis was conducted using SPSS version 24.0.

**Results:** A total of 775 participants completed the survey. Among this final sample, average age was 24 years. Among the survey participants, 455 (59%) were men, 444 (had bachelor’s degree, 364(47%) had monthly income between Pakistani Rupees (PKR) 25,000 to 60,000, and 483 (62%) were married. The overall correct response rate of knowledge was 73.2%. The score in COVID-19 negative attitude was highest. 455 males reported handwashing. Majority (89%) had prior knowledge regarding COVID-19 clinical symptoms. The 71% showed their confidence in ability for Pakistan to control COVID-19 outbreak. 87% avoided reducing outdoor activities.

**Conclusion:** The study findings suggest that residents of Islamabad city have adequate levels of knowledge regarding COVID-19. However, it is paramount to introduce awareness campaigns and public health education to promote the knowledge and practice of COVID-19 disease.

## Background

Coronavirus Disease or Covid-19 was first reported by World Health Organization in Wuhan, Hubei province, China in 31^st^ December 2019 (1). On 30^th^ January, 2020 of WHO to declare COVID-19 a Public Health Emergency of International Concern due to its rapid spread(2). WHO declared this disease as pandemic after the disease spread to over 110 countries and territories including Pakistan(3).

COVID-19 is a highly contagious disease that is caused by SARS-CoV-2. The SARS-CoV-2 belongs to wide range of beta coronavirus family and is closely related SARS-CoV and MERS-CoV(4, 5). It is an RNA virus which has single stranded RNA positive sense RNA genome. It has been believed that SARS-CoV-2 initially infected human host through intermediate host such as wild animal species handled by humans. The major clinical symptoms of COVID-19 include dry cough, fever, myalgia, difficulty in breathing and fatigue, other reported symptom may include the loss of taste and smell while some reported diarrhea, congestion and running nose (6). Close contact with infected person upon sneezing or coughing can cause spread of COVID-19.

The virus has an incubation period of 4 to 15 days. Individuals with medical condition like cancer, diabetes and cardiovascular disease and elderly people have higher chances to contract severe infection (7). Several vaccines are being used to prevent and control COVID-19 and its ongoing variants in Pakistan (8). On the 26 February 2020, Pakistan reported its first confirmed case who had recently travel from Iran. Till 15 December 2022, there has been 1.5 million cases with 30,000 reported fatalities with highest in Sindh Province (n= 595,372) followed by Punjab province (n= 523,226), Khyber Pakhtunkhwa province (n= 224,797), Islamabad (n= 139,641), Azad Jammu Kashmir (n= 44,336) Baluchistan (n=36,027) and Gilgit Baltistan (n= 12,087) (9). COVID-19 pandemic has affected Islamabad City, the capital of Pakistan. Initially several prevention methods were adopted to control virus including temporary suspension of local transport, ban on public gathering and establishment of quarantine centers while the state moved towards more manageable lockdown called “smart lockdown”(10). The citizens of Islamabad city were instructed to stay home and keeping social distance with others. According to the Knowledge, Attitudes and Practices (KAP) theory, the success of these preventive is influenced by KAP (11). Hence, the aim of this study is to identify the current situation on KAP regarding COVID-19 in working professionals in order to inform the control strategies in context of COVID-19 pandemic.

## Methodology

### Participants

The survey study was conducted between 5^th^ to 19^th^ of May 2020, which took place during smart lockdown in Islamabad. Employees of several government institution, National Institute of Health, were instructed to observe physical presence with strict observation of COVID-19 Standard Operating Procedures (SOPs). Hence, the data was collected in person.

This in-person survey was carried out in various offices of NIH and affiliated institutions through the diverse network of author’s colleagues and peers, residing in Islamabad city. The survey consisted of three pages of questionnaire document taken with in-person interviews. The questionnaire was thoroughly discussed and evaluated by a team that consisted of three members. The team was trained to conduct physical interviews observing strict COVID-19 SOPs. The interviewers briefly introduced the survey and described its objectives, declaration of confidentiality. All participants were Pakistani nationals, with the age of 18 or above, understood the survey, and agreed to participate in the study. The team ensured that only residents of Islamabad city were interviewed to fill the survey.

The ethical committee of NIH approved this cross-sectional survey’s procedures and protocols of informed consent prior to beginning of interview. A choice of ‘yes’ or ‘no’ was given to participants to confirm their willingness of participation. Only after the affirmative response from the participant the interview was conducted.

The Center for Epidemiology Study Depression Scale is used to develop COVID-19 Negative Affective Attitude Scale. The Likert five-point scoring method is adopted for this scale. Negative feeling in this dimension is indicated by higher score on the scale. The higher than 0.90 score at Kaiser Meyer Olkin (KMO) test as well as Bartlett’s score reached at significant level indicates suitability for factor analysis and the scale incorporated common factors. After the extraction, by exploratory factor analysis (EFA) with rotation by varimax and principal component, **(Table 3)** illustrates factor leading, explained variations, correlation commonality and reliability, with overall reliability of 0.88, showing internal consistency.

### Measures

The interview consisted of two parts such as KAP and sociodemographic. Gender, age, marital status, occupation, education, monthly household income, and current residence were the sociodemographic variables of this study. During the previously mentioned period, 766 participants were successfully interviewed.

The survey was designed by the authors using strict social distancing, mask wearing, and disease management of COVID-19 guidelines issued under the authority of Ministry of Health Services Regulation and Coordination, Pakistan. The KAP part of interview was furthered divided two three sections: Knowledge Aptitude and Practice. The knowledge section has 13 questions: K1 to K5 are clinical presentations, K6 to K11 are about prevention, transmission, and control, K12 is about isolation and K13 is about COVID-19 vaccination. The questions in Knowledge section were answered with option: ‘True’ or ‘False’ or ‘I don’t know’. Valid response was assigned with 1 mark and 0 mark for invalid or unknown response. The overall range of knowledge score was between 0 to 14, where higher score indicates better knowledge of COVID-19. The coefficient of Cronbach’s alpha of knowledge was 0.71 in our sample that shows reasonable internal consistency.

### Statistical Analysis

The frequencies of correct knowledge answers and various attitude and practice have been described. Knowledge scores, attitude and practice of different individuals based on demographic characteristics were compared with independent sample t test, Chi square test, or one way analysis variance. The SPSS version 21.0 was used to conduct data analysis. The statistical significance level was set up at p < 0.05, (two sided).

## Results

A total of 775 participants successfully completed KAP survey. Maximum number of participants were male 59% (n=455) than female 41% (n=320) with 36% (n=277) of them aged between 18 to 30 years (median: 24).

About 57% (n=444) participants had bachelor’s degree, and 47% (n=364) had monthly household income ranging from PKR 25,000 to PKR 60,000, as shown in. A total of 62% (n=483) participants were married, and 68% (n=526) participants were urban residents **(Table 1)**.

**Table 1.** Descriptive statistics of survey respondents (n = 775)

| Sociodemographic characteristics | Total (n=775) |  |
| --- | --- | --- |
|  | n | % |
| **Gender** |  |  |
| Male | 455 | 59 |
| Female | 320 | 41 |
| **Age-group (years)** |  |  |
| 18-30 | 277 | 36 |
| 31-40 | 213 | 27 |
| 41-50 | 133 | 17 |
| 51-60 | 103 | 13 |
| 60+ | 49 | 6 |
| **Education** |  |  |
| High school | 77 | 10 |
| Bachelor's degree | 444 | 57 |
| Graduate/professional degree | 254 | 33 |
| **Monthly household income in PKR** |  |  |
| < 25000 | 53 | 7 |
| 25000-60000 | 364 | 47 |
| 60000-100000 | 257 | 33 |
| > 100000 | 101 | 13 |
| **Marital Status** |  |  |
| Married | 483 | 62 |
| Unmarried | 263 | 34 |
| Others^ | 29 | 4 |
| **Residence** |  |  |
| Urban | 526 | 68 |
| Rural | 249 | 32 |

The overall average of correct answers for Knowledge section of interview for the survey was 72.3%, wherein the subsection (K10) ‘handwashing, social distancing and staying at home as effective preventive measures’ had the highest percentage (97%) of correct responses. Whereas (A15) of attitude section regarding smart ‘lockdown strategy’ can curb COVID-19 in Pakistan, had 71% correct answer. In practice section, 87% answered correctly about reducing outdoor activity during pandemic **(table 2)**.

**Table 2.** Descriptive statistics of KAP questionnaire survey respondents (n = 775)

| Questions | Options |
| --- | --- |
| **Knowledge (correct rate, % of the total sample)** |  |
| **K1:** The main clinical symptoms of COVID-19 are fever, cough, and breathing difficulty. (89%) | True, False, Don't know |
| **K2:** Do you think runny nose and sneezing are also symptoms of COVID-19 virus. (44%) | True, False, Don't know |
| **K3:** Do you think early deduction and supportive treatment can recover patients from the infection? (72%) | True, False, Don't know |
| **K4:** Persons with COVID-19 cannot infect to others when fever is not present. (12%) | True, False, Don't know |
| **K5:** The COVID-19 spread through coughing, sneezing and touching contaminated objects. (93%) | True, False, Don't know |
| **K6:** Surgical mask could be used to prevent COVID-19? (90%) | True, False, Don't know |
| **K7:** To prevent the infection, individuals should avoid going to crowded places such as train stations, shopping malls and avoid taking public transportations. (83%) | True, False, Don't know |
| **K8:** It is not necessary for children and young adults to take measures to prevent the infection by COVID-19. (25%) | True, False, Don't know |
| **K9:** Isolation (social distancing and stay at home) is an effective way to reduce the spread of the virus. (88%) | True, False, Don't know |
| **K10:** The COVID-19 will be controlled by practicing hand washing, social distancing, and staying at home. (97%) | True, False, Don't know |
| **K11:** The COVID-19 will be controlled by avoiding travelling across/ within country during this outbreak. (91%) | True, False, Don't know |
| **K12:** People having contact with COVID-19 patient should be immediately isolated in a proper place. (87%) | True, False, Don't know |
| **K13:** The COVID-19 vaccine is available against this virus. (7%) | True, False, Don't know |
| **Attitude** |  |
| **A14:** Do you agree that lockdown strategy can finally controlled the spread of COVID-19 virus? (41%) | Agree, Disagree,<br>Don't know |
| **A15:** Do you have confidence that Pakistan can win the battle against the COVID-19 virus? (71%) | Agree, Disagree,<br>Don't know |
| **Practice** |  |
| **P16:** In recent days, have you gone to any crowded (social or religious) place? (69%) | Yes, No |
| **P17:** If you have fever or cough, would you like to isolate/quarantine yourself? (78%) | Yes, No |
| **P18:** During COVID-19 outbreak, have you reduced the outdoor activities? (87%) | Yes, No |

The participants’ negative attitude towards themselves and life indicates a low prevalence of these attitude during survey period **(table 3)** as well as with negative scores which are significantly higher in attitude (*p*=0.002) towards self than attitude towards life, indicates greater effect of external living environment brought about by the pandemic than the individuals’ inner psychology. Participants that have expressed preventive behavior scored higher in hygiene habits than in helping others to prevent COVID-19 spread and reducing outdoor activities which indicates that participants were generally more willing to change their overall COVID-19 behavior and hygiene habits **(Table 4)**.

**Table 3.** COVID-19 negative affective attitude scale.

| Characteristics | Commonality | Factor Analysis |  | Explained Variations | Reliability |
| --- | --- | --- | --- | --- | --- |
|  |  | Negative Self-Perception | Negative Perception of Life |  |  |
| 1-I feel depressed. | 0.81 | 0.84 | 0.12 | 46.79 | 0.91 |
| 2-I think my life is useless. | 0.69 | 0.75 | 0.19 |  |  |
| 3-I feel I will never be able to travel anywhere anymore | 0.56 | 0.69 | 0.07 |  |  |
| 4-I feel alone | 0.59 | 0.8 | 0.09 |  |  |
| 5-Feeling isolated and stressed | 0.83 | 0.81 | 0.17 |  |  |
| 6-I feel it's never-ending thing | 0.67 | 0.73 | 0.33 |  |  |
| 7-I feel I am wasting my time | 0.52 | 0.67 | 0.4 |  |  |
| Reverse |  |  |  |  |  |
| 7-I utilize my time in best way | 0.77 | 0.17 | 0.88 | 16.23 | 0.76 |
| 8-I spent time with family (accompanied) | 0.81 | 0.21 | 0.89 |  |  |
| 9- I think positive about future | 0.58 | 0.19 | 0.76 |  |  |

Similarly, the test higher than 0.90 score at KMO test and Bartlett’s score reached at significant level indicates suitability for factor analysis and scale incorporated common factors for COVID-19 Preventive Behavior Scale as well. Three factors were extracted using principal component analysis and EFA with rotation using varimax: (1) ‘willingness to integrate pandemic prevention and hygiene measures into practice, comply with preventive method regulated by state and schools; (2) ‘cooperation with social distancing to prevent direct contact’; (3) ‘willingness to participate in society by demonstrating altruistic attitude’. The Likert five-point scoring method is adopted for this scale as well. A more proactive response towards COVID-19 is indicated by higher score on this scale. This table shows reliability, explained variation, factor loading, and correlation commonality **(Table 4)**.

**Table 4.**
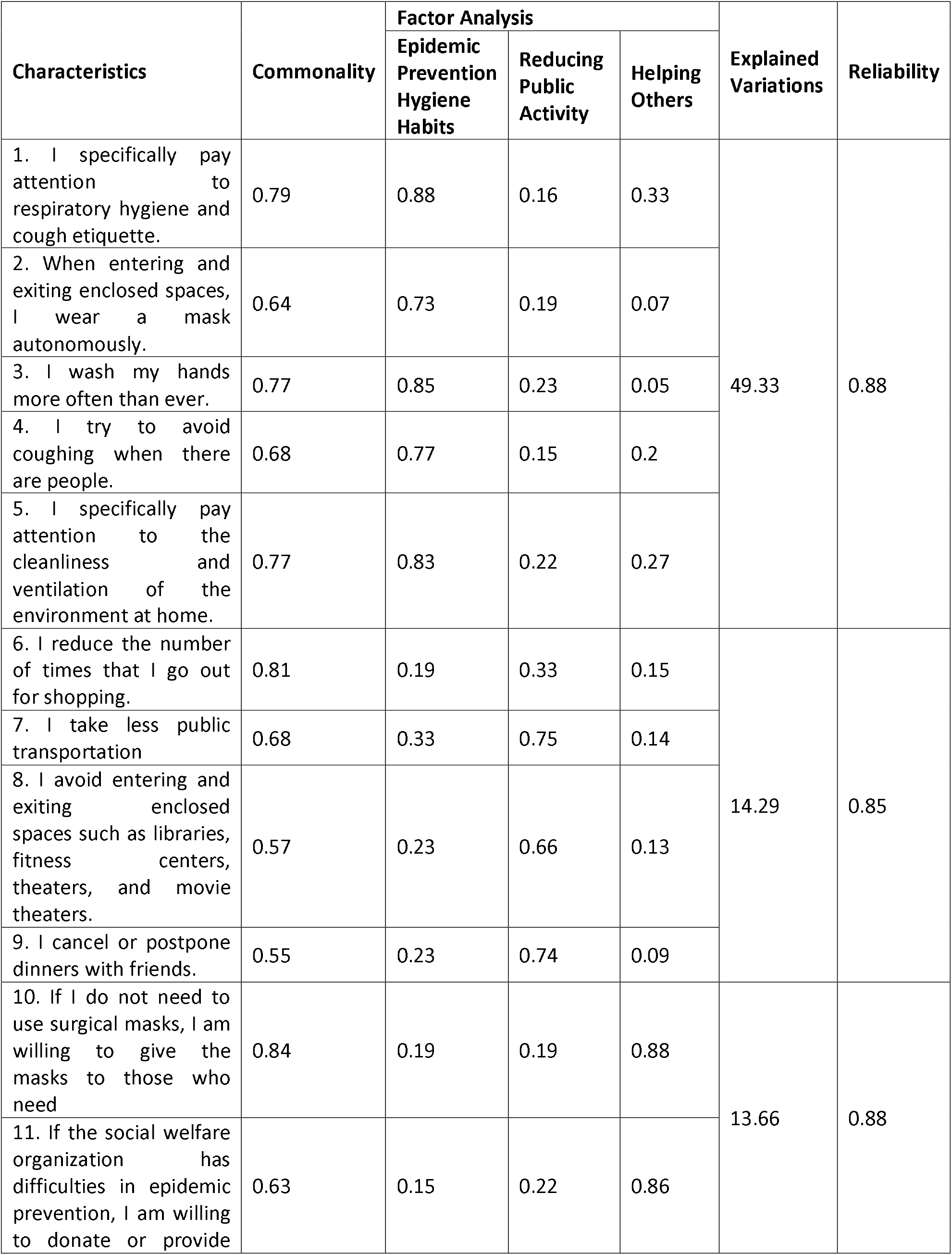
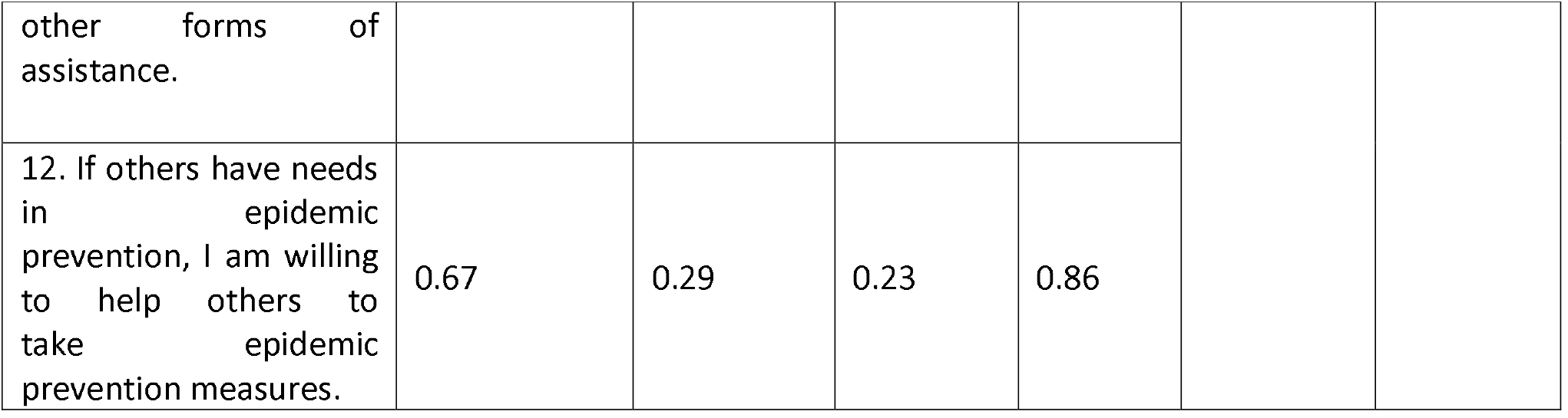
COVID-19 preventive behavior scale.

Approximately 40.4% (n=313) male participants have reported that they avoided crowded place verses 28.2% (n=249) female participants. While the 29.0 % (n=225) males ensured social distance as compared to female 28% (n= 219). A greater number of participants reported handwashing with 57.4 % (n= 455) males and 41.3% (n=320) females. Furthermore, 38.8% (n=301) males and 30.5% (n=237) females reported feeling of isolation and stress **(Figure 1)**.

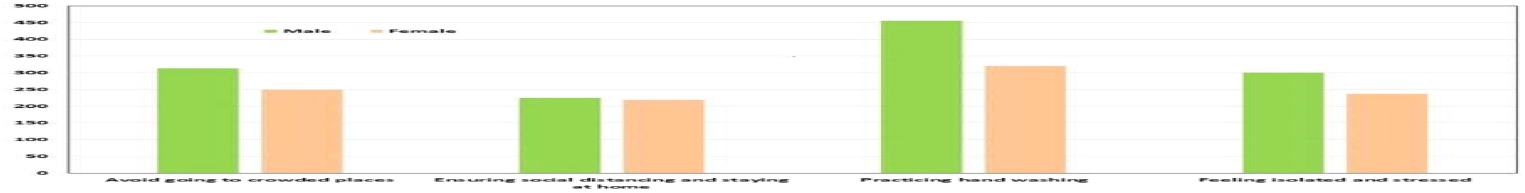

Most participants, 89% (n=690), had prior knowledge regarding clinical symptoms such as fever, cough, and breathing difficulty. While 59% (n=457) of participants disagreed that COVID-19 infected person cannot infect others when fever is not present. The 72% (n=558) of participant believed that a person could recover from infection with early diagnosis and supportive treatment. Furthermore, 90% (n=697) participant agreed that surgical mask can be used to prevent COVID-19 **(Figure 2)**.

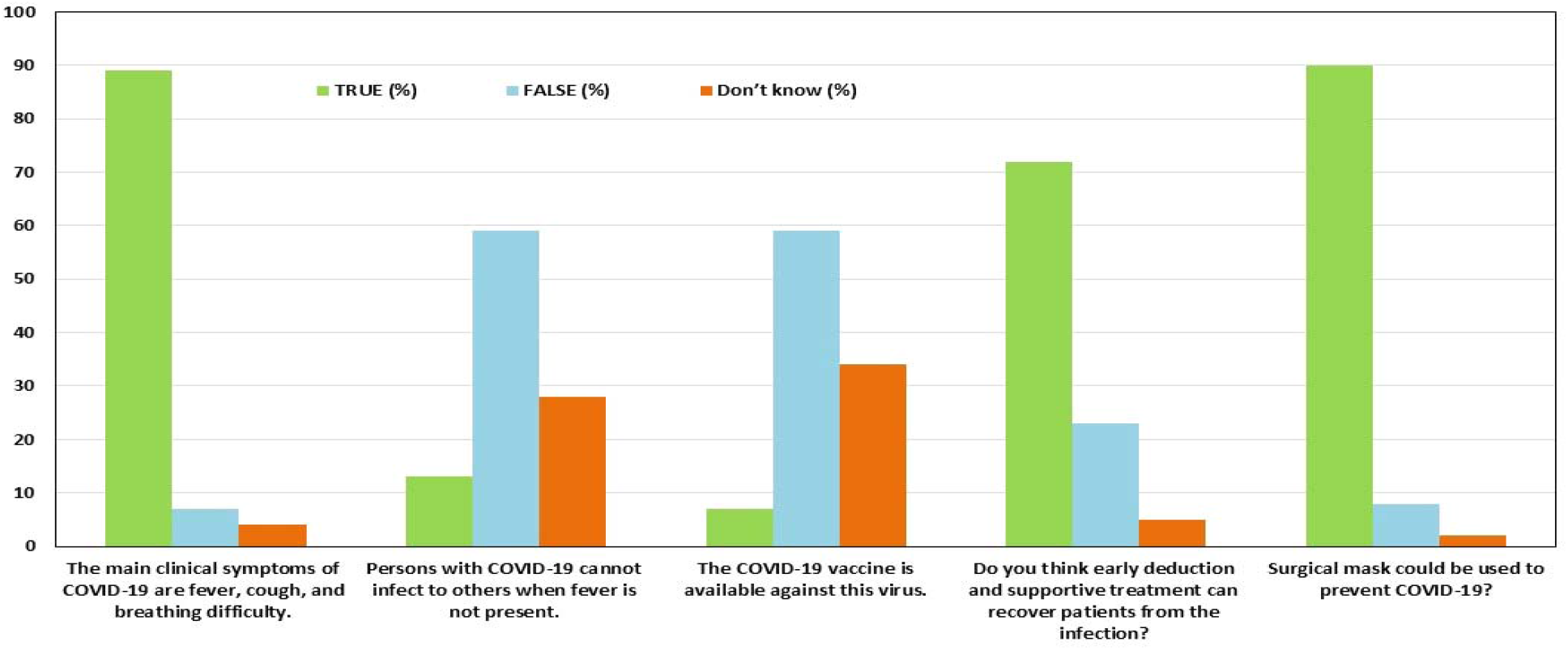

The 50% (n=387) of participants agreed that lockdown strategy will effectively control the transmission of COVID-19. Meanwhile, 71% (n=550) of participants showed their confidence that Pakistan could control COVID-19 disease.

## Discussion

The lack of knowledge and awareness as well as diverse public opinion about COVID-19 facilitates its progression in vulnerable population (12). There has been scarcity of published survey studies on public knowledge, attitude, and practice towards COVID-19 in Pakistan. Bulk of uncertainty and misinformation have necessitated public health organizations to surveil public opinion and practice and implement pertinent pandemic management strategies. This study indicates diversification in public beliefs and opinion towards COVID-19 (13). KAP analysis was made by data collected from interviewing respondent of Islamabad.

Most of young adult responded and agreed to our survey because young adults are more open minded about COVID-19 pandemic. Our study also indicates that the most study participant had sufficient knowledge about COVID-19 because the number of correct responses of COVID-19 was high enough. A study from Kingdom of Saudi Arabia supports the finding of this study that represents correct answer rate of 81.64% (14).

The participants had relatively negative attitude and feelings towards their personal external life during pandemic which is primarily because of several inconveniences in life caused by COVID-19 pandemic such as travel ban, reduced outdoor activities and socializing, and work pressure. The increase in negative perception of life and decline in emotional wellbeing should receive more attention especially in younger groups which may experience typical stress of adolescence growth as well as stress related to COVID-19 pandemic restrictions (15).

At the time when this survey was conducted and interviews were being done, most people had the exposure to basic knowledge about the disease via electronic and print media. Government of Pakistan (GoP) along with NIH launched public awareness campaign which helped to improve knowledge of Pakistani population regarding COVID-19 (16). The studies conducted in Pakistan and China has shown that public awareness campaigns against COVID-19 improve public knowledge attitude and practice (17, 18). Most candidates were aware of basic preventive measurements such as social distancing, handwashing and staying home which convinced them that following these measures will prevent the spread of COVID-19. These measures which we evaluated from responses of candidates were based on WHO guidelines for prevention and control for COVID-19.

Regarding COVID-19 pandemic, participants have shown positive attitude, most of participants believe that effective measures can successfully control COVID-19. This optimistic approach towards pandemic may have developed by partial lockdown strategies like Smart lockdown that eased several harsh restrictions such as travel ban, closure of nationwide educational institutes as well as non-essential markets (19). Hence, our finding also emphasizes higher degree of trust, knowledge, and reasonable attitude towards government interventions taken during pandemic emergences, which is also similar to another Chinese study (20).

This survey also indicates that 69 % of participants practiced appropriate preventive measurements such as avoiding public crowd, most commonly religious events while most people reported reduced outdoor activities

## Conclusion

This study thoroughly provides information regarding KAP of COVID-19. The outcome of this study indicates reasonable level of knowledge of Islamabad city residents on COVID-19. Furthermore, it is pertinent to provide awareness campaign and adequate public health education to improve the knowledge attitude and practice towards COVID-19, to curb COVID-19 transmission.

### Limitations

We were unable to reach out to more group of people which may have insufficient education, elderly, more general public which might have made our study more diverse in outcomes.

The interviews were conducted on NIH premises and related offices which uneducated and elderly people. Hence, the study covers responses which are mostly young adults who are familiar with knowledge regarding COVID-19

## Data Availability

All data produced in the present work are contained in the manuscript

## Data Availability

The author confirms that all data generated or analysed during this study are included in this published article.

## Ethics Approval Statement

The study was performed in accordance with the COVID-19 Pandemic: Guidelines for Ethical Healthcare Decision-Making in Pakistan.

## Informed Consent to participate statement

Verbal informed consent was obtained prior to the interview.

“Informed consent was obtained from all individual participants included in the study. The Ethical Committee of National Institute of Health (NIH) Islamabad, approved this cross-sectional survey’s procedures and protocols of informed consent prior to beginning of interview. Participants were informed about the purpose of the study, the procedures involved, and their right to withdraw from the study at any time without penalty. A choice of ‘yes’ or ‘no’ was given to participants to confirm their willingness of participation. Only after the affirmative response from the participant the interview was conducted. Participants’ identities have been anonymized to ensure confidentiality and their responses were kept confidential and used solely for research purposes.”

